# Research on Epilepsy Detection and Recognition Based on the Combination of Time Frequency Transform and Deep Learning Model

**DOI:** 10.1101/2025.11.02.25339347

**Authors:** Canhui Wang, Yan Li, Haoran Tang, Zongfang Ren, Tianqi Xu

## Abstract

To improve the detection performance of epileptic electroencephalogram (EEG) signals and address their non-stationary characteristics, this paper compares the combined effects of continuous wavelet transform (CWT), short-time Fourier transform (STFT), along with three neural network models—EEGNet, AlexNet, and Shallow ConvNet—and incorporates innovative designs. Specifically, Focal Loss, dynamic data augmentation, and an early stopping mechanism are introduced at the training stage to enhance the model robustness. Additionally, EEGNet is optimized by integrating an SE (Squeeze-and-Excitation) attention module, improving depthwise separable convolution (where a (3,16) kernel is used in the first layer), and dynamically adapting dimensions to reduce errors. For Shallow ConvNet, improvements are made by adopting layered convolution to extract “time-frequency” features and average pooling to adapt to long data blocks. The results show that the recall rate of the CWT+Shallow ConvNet combination reaches 100% with an accuracy of 99.14%, while the accuracy of the CWT+EEGNet combination achieves 100%. These findings verify the effectiveness of combining precise time-frequency features with optimized models, providing support for clinical practice.

## Introduction

Electroencephalography (EEG), as a non-invasive detection technology, amplifies and converts the bioelectric activity generated by brain neurons into analyzable EEG curves by placing electrodes on the scalp. It not only plays a core role in the clinical diagnosis of epilepsy, cerebrovascular diseases, and other conditions, but also serves as key technical support in the field of brain-computer interface (BCI)**^Error! Reference source not found.^**. By decoding features such as motor imagery^[1]^ and steady-state visual evoked potentials, EEG enables direct communication between the human brain and external devices, and has demonstrated significant value in scenarios such as disability assistance and rehabilitation. In recent years, the integration of EEG signals with deep learning has further driven improvements in BCI performance^[3]^, yet it still faces challenges including low signal-to-noise ratio and substantial individual differences. In the future, combining multimodal fusion with adaptive algorithms^[4]^ will be a crucial breakthrough direction.

The precise detection of epileptic EEG signals is in urgent demand in clinical practice. Its core goal is to provide an objective basis for the diagnosis, classification, treatment evaluation, and prognosis of epilepsy by capturing in real time and identifying accurately abnormal discharges (such as spike waves and sharp waves) in electroencephalograms (EEG)^[5]^. For pediatric patients, their EEG signals are characterized by abundant high-frequency rhythms, diverse seizure patterns, and high susceptibility to developmental stages. Consequently, traditional manual interpretation is not only time-consuming but also suffers from issues such as high missed detection rates and strong subjectivity—problems that directly affect the timing of early intervention^[6]^. Additionally, patients with drug-resistant epilepsy require long-term EEG monitoring to localize epileptic foci^[7]^, thereby providing targets for surgical treatment^[8]^. However, existing technologies lack sensitivity in identifying weak abnormal discharges during seizures, causing some patients to miss the opportunity for surgery. Therefore, there is an urgent need for automated detection technologies in clinical practice to achieve high recall and accuracy in the detection of epileptic EEG signals. This is particularly critical for pediatric epilepsy screening, long-term monitoring, and preoperative evaluation^[9]^, as it can improve diagnostic efficiency, optimize treatment plans, and ultimately enhance patients’ quality of life.

This study conducts a comparative investigation on the detection task of epileptic electroencephalogram (EEG) signals, based on multiple deep learning models and time-frequency feature extraction methods. The main research content includes the following: adopting Shallow ConvNet, EEGNet, and AlexNet as the core model architectures; applying Continuous Wavelet Transform (CWT) and Short-Time Fourier Transform (STFT) to perform time-frequency feature transformation on EEG signals; constructing multiple classification models; and systematically evaluating their performance. In the experiments, the stability of the models was verified via 5-fold cross-validation, with a focus on comparing the accuracy, precision, and recall of different models under the input of CWT-derived and STFT-derived features. Among these models, the combination of Shallow ConvNet and CWT exhibited superior performance in terms of accuracy and recall, while the combination of EEGNet and CWT achieved optimal performance in accuracy.

In summary, the main contributions of this study are as follows:

(1) Optimization design for training: Based on the characteristics of epileptic EEG signals, the Focal Loss function, a dynamic data augmentation strategy, and an early stopping mechanism are introduced during model training to enhance the model’s robustness in recognizing epileptic samples.
(2) Modifications to the classic EEGNet model: Three key modifications are proposed: first, an SE (Squeeze-and-Excitation) attention module is integrated to strengthen the model’s focus on the salient features of epileptic EEG signals through adaptive channel weight assignment; second, depthwise separable convolution is optimized by using (3, 16) kernels in the first layer to capture local time-frequency patterns, which reduces redundant parameters in deep convolutional layers, and activation functions are applied to mitigate training fluctuations caused by the sparsity of EEG data; third, dynamic dimension adaptation is adopted to automatically calculate the input dimension of the fully connected layer based on the feature sizes of CWT/STFT, thus avoiding feature truncation or padding errors caused by fixed dimensions.
(3) Improvements to the Shallow ConvNet model: The kernel in the first layer captures time-series information, while the deep convolution in the second layer covers the entire frequency scale, enabling hierarchical extraction of “time-frequency” features; average pooling is used to downsample the time dimension, which preserves information from key time points while reducing computational complexity and adapting to the global feature learning of 10-second-long data blocks.

### Related Work

The integration of time-frequency transformation and deep learning provides a powerful tool for analyzing EEG signals: time-frequency transformation can convert non-stationary EEG signals into two-dimensional feature maps that contain time-frequency correlation information, thereby effectively capturing the time-frequency distribution of transient events (e.g., spikes) in epileptic EEG signals^[10]^. This enables deep learning models to automatically mine discriminative information from the feature maps, avoiding the limitations of manual feature design and significantly improving classification accuracy^[11]^. However, the field still faces key challenges: first, the empirical setting of time-frequency transformation parameters may lead to feature redundancy or the loss of critical information; second, deep learning models are sensitive to data distribution, making them prone to overfitting in scenarios involving small-sample pediatric EEG data; additionally, the “black-box” nature of model decision-making^[12]^ conflicts with the clinical demand for interpretability, which limits the models’ application in critical scenarios such as epilepsy diagnosis.

Current research on time-frequency analysis and model adaptation for EEG signals has three significant limitations: first, the setting of time-frequency transformation parameters lacks specificity—most studies adopt a universal scale range (e.g., the default 3–30 for CWT) without considering the specificity of the high-frequency dominant band (10–20 Hz) in pediatric EEG. This results in limited accuracy when extracting transient features such as spike waves^[13]^; second, there is a lack of collaborative optimization between models and features. Although existing studies have verified the compatibility of CWT and CNNs, there is no systematic comparison of the balance between recall and accuracy across different lightweight models (e.g., Shallow ConvNet and EEGNet), making it difficult to meet the clinical requirement of “prioritizing missed detection prevention”; third, the utilization of multi-channel spatial features is insufficient. Most existing methods simply concatenate multi-channel time-frequency maps for model input, ignoring the cross-channel propagation pattern of abnormal discharges.

Zare et al^.[14]^ employed a Support Vector Machine (SVM) classifier for classification tasks. The results showed that the OMP-based technique achieved an average specificity of 96.58%, an average accuracy of 97%, and an average sensitivity of 97.08% across different classification tasks; while the DWT-based technique performed better, with an average sensitivity of 99.39%, an average accuracy of 99.63%, and an average specificity of 99.72%.Tara et al.^[15]^ proposed a hybrid Random Forest-Convolutional Neural Network (RF-CNN) model. This model integrates a feature-based Random Forest (RF) machine learning model with an image-based Convolutional Neural Network (CNN) deep learning approach, addressing the limitations of previous models that relied solely on either feature-based machine learning or image-based deep learning alone.Malakouti et al.^[16]^ proposed a lightweight and interpretable framework for epileptic seizure detection. The core of this framework lies in combining Discrete Wavelet Transform (DWT) with bandpass filtering to extract robust time-frequency features from EEG signals, which effectively reduces the interference of the nonstationarity and noise of EEG signals on subsequent classification tasks.Zhao, et al.^[17]^ proposed a novel EEG-based epileptic seizure detection method integrating time-frequency features. The proposed ConvNeXt-SimAM model exhibited excellent performance: Accuracy at 98.83%, Specificity at 97.68%, Sensitivity at 96.86%, and Kappa score at 0.9551.Liu, Yingjian, et al.^[18]^ demonstrated that the phase information in EEG signals is useful for epileptic seizure detection. On the CHB - MIT database, when the phase input was additionally provided to the CNN model, the AUC-ROC of detection increased by 6.68%. Through the post - processing of channel fusion for the output of the CNN model, the model achieved a sensitivity of 79.59% and a specificity of 92.23%, surpassing some existing methods. The usefulness of phase inputs in the CHB - MIT and Bonn databases was verified.Abdulwahhab et al.^[19]^ proposed a concerted deep machine learning model that integrates two simultaneous techniques for epileptic seizure detection. When using CWT scalograms as the input for the CNN, the model achieved an accuracy of 99.57% on both the Bonn University dataset and the CHB-MIT dataset. When using STFT spectrograms as the input for the CNN, it reached an accuracy of 99.26% on the Bonn University dataset and 97.12% on the CHB-MIT dataset.Islam et al.^[20]^ proposed a novel framework capable of detecting epilepsy from noisy EEG signals and innovatively conducted age-stratified analysis. When models that performed well in the general analysis were applied to two age groups, the results showed that the group under 40 years old achieved significantly higher detection accuracy, revealing age-related differences in EEG-based epilepsy detection.

In summary, although researchers at home and abroad have conducted extensive studies on EEG-based epileptic seizure detection and recognition and achieved considerable results, research on analyzing epileptic EEG signals through the integration of time-frequency transformation and deep learning remains relatively limited. Furthermore, existing methods suffer from drawbacks such as weak anti-interference capability, insensitivity to local features, and dependence on specific electrode layouts. Therefore, developing a universal and efficient method for epileptic seizure detection and recognition is of great significance for ensuring the accuracy of epilepsy diagnosis in clinical practice.

## 3 Design and Implementation

### 3.1 Experimental data and environment

The dataset used in this experiment is from the CHB-MIT Scalp EEG Database. Collected by Boston Children’ s Hospital in 2010, the CHB-MIT scalp EEG data is stored in the.edf (European Data Format) file format and publicly available on the PhysioNet platform. This dataset includes scalp EEG signal recordings from 23 pediatric epilepsy patients, consisting of 5 males (aged 3–22 years) and 17 females (aged 1.5–19 years). All EEG data were collected using the 10–20 International System of Electrode Placement, and most files contain 23 EEG channels with a sampling frequency of 256 Hz and a resolution of 16 bits.

The computing device used in this experiment is a computer equipped with an NVIDIA GeForce RTX 4060 GPU. Based on the Ampere architecture, this GPU has 8 GB of GDDR6 memory and supports CUDA 11.8-accelerated computing. The model training and inference processes were implemented using the PyTorch framework (version 2.0.1)—this framework invokes CUDA cores for parallel computing, significantly improving computational efficiency. The computer is also configured with an Intel Core i7-12650H CPU and 40 GB of DDR4 memory, which ensure efficient data preprocessing and batch data loading. The training parameters are set as follows:

1. Optimizer: AdamW (initial learning rate = 5e-4, weight decay = 1e-3);
2. Loss function: Focal Loss (α = 0.6, γ = 2.0), designed to mitigate class imbalance;
3. Training strategy: 5-fold cross-validation was adopted, with 20 epochs per fold.

An early stopping mechanism was added—training for a given fold terminates if no performance improvement is observed after 8 consecutive epochs.

### 3.2 Characteristics and Preprocessing of Epilepsy EEG Signals

Feature extraction and preprocessing of epileptic EEG signals are core steps for achieving automatic epileptic detection, as they directly influence model performance. EEG signals during epileptic seizures exhibit significant physiological specificity, and the goal of preprocessing is to preserve these key features while removing noise interference—laying a foundation for subsequent analysis and modeling. Abnormal discharges in epileptic EEG signals primarily originate from the synchronous firing of neurons, which manifest as the following typical features:

(1) Waveform features: Spike waves, sharp waves, spike-and-slow complex waves, and high-amplitude, high-frequency paroxysmal rhythms during both ictal (seizure) and interictal (interseizure) periods.
(2) Quantitative features: In the time domain, the amplitude and variance of the signal increase significantly; in the frequency domain, the energy of the gamma band rises sharply while spectral entropy decreases; in the time-frequency domain, transient energy bursts occur, and the frequency may evolve from high to low as the seizure progresses.

The purpose of preprocessing is to preserve abnormal features while removing noise. Based on the typical features of epileptic EEG signals, we adopted the following corresponding preprocessing steps:

(1) Noise filtering: A 1–50 Hz band-pass filter was used to retain key frequency bands, effectively filtering out DC drift and high-frequency electromyographic (EMG) noise.
(2) Signal segmentation: Long-duration EEG signals were divided into fixed-length sub-blocks of 5–10 seconds. For edge segments with insufficient length, zero-padding was applied to balance feature integrity and computational efficiency.
(3) Time-frequency transformation and normalization: Time-domain signals were converted into two-dimensional time-frequency features using either CWT or STFT, which highlights transient anomalies; Z-score normalization was adopted to eliminate amplitude differences and prevent noise from interfering with model performance.
(4) Sample balancing and augmentation: A sample ratio of 2:1 (normal to abnormal) was maintained by undersampling normal samples and oversampling abnormal samples via the SMOTE (Synthetic Minority Oversampling Technique) algorithm; to expand the dataset and enhance model robustness, noise addition or amplitude scaling was applied to abnormal samples, while time shifting was performed on normal samples.

### 3.3 Method Design and Implementation

This section describes the system employed in this study, which is designed for the automatic identification and detection of key patterns in epileptic EEG signals, as illustrated in Figure 1. The system primarily comprises four modules: a data preprocessing module, a model construction module, a training optimization module, and an epilepsy detection and evaluation module. Detailed descriptions of these modules are provided as follows:

(1) Signal Preprocessing Module: The function of this module is to convert raw electroencephalogram (EEG) signals into input-compatible signals for deep learning models while highlighting their physiological relevance. First, this module applies a 4th-order Butterworth band-pass filter to perform 1–50 Hz band-pass filtering, which removes DC components and high-frequency noise. Next, the filtered data is segmented into blocks with a duration of 10 seconds per block. Then, continuous wavelet transform (CWT) is used to convert the time-domain signals into two-dimensional time-frequency maps (frequency × time), thereby preserving the non-stationary characteristics of EEG signals. To enhance the model’s robustness to signal variations, Gaussian noise, time shifts, and amplitude scaling are introduced into the training set. Finally, mean-standard deviation normalization is applied to the time-frequency maps to eliminate the impact of inter-individual amplitude differences. The core transfer function of the Butterworth band-pass filter is as follows^[21]^:

**Figure 1.**
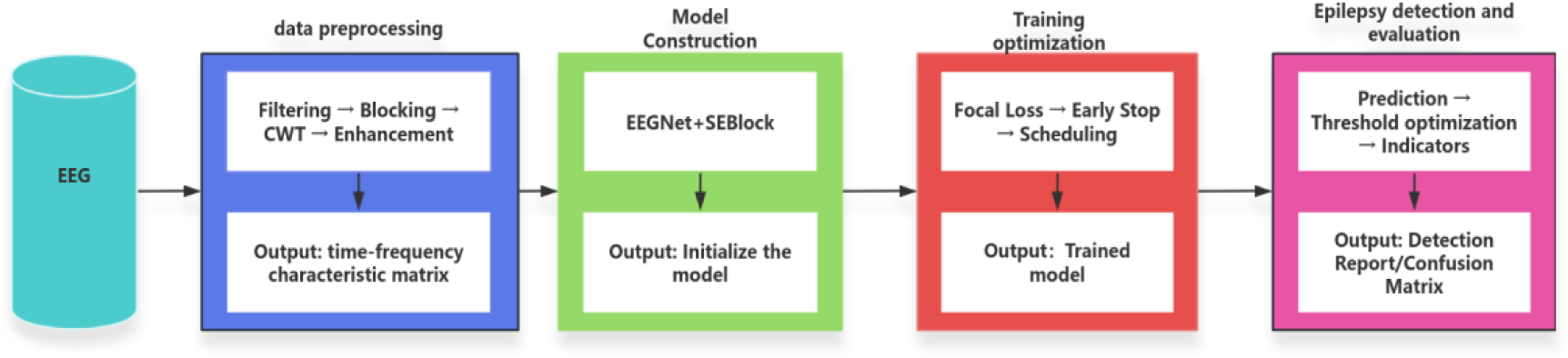
Flow Chart of Epilepsy Automatic Recognition and Detection System

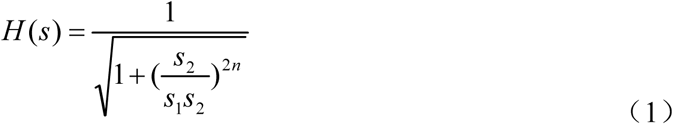

Where *H(s)* denotes the transfer function of the filter, and *s* is the complex frequency variable; N represents the order of the filter; *s_1_* and *s_2_*are the cutoff frequencies of the band-pass filter.

(2) Model Construction Module: The function of this module is to design a deep learning model adapted to EEG time-frequency features and realize the classification of normal and epileptic signals. The experimental training model adopts EEGNet. Given the limited sample size, depthwise separable convolution is used to reduce the number of parameters. By embedding the SEBlock (Squeeze-and-Excitation Block) module to dynamically adjust feature weights and focus on key time-frequency regions (e.g., high-frequency features during epileptic seizures), regularization of the model is achieved through Dropout and BatchNorm (Batch Normalization) to suppress overfitting and improve its generalization ability. The BatchNorm operation—critical for stabilizing training—is defined as follows^[22]^:

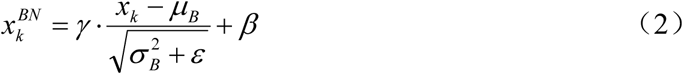

where *μ_B_*and *σ_B_^2^* are the mean and variance of the batch, respectively; *γ*and *β*are learnable parameters; and *ɛ* is a small constant added to avoid division by zero.

For the design of the Shallow ConvNet model: the kernel in the first convolutional layer captures time-series information, while the deep convolution in the second layer covers the entire frequency scale—enabling hierarchical extraction of “time-frequency” features. The output size of its convolutional layers (a factor that determines the dimension of feature maps) follows the formula below**^Error! Reference source not found.^**:

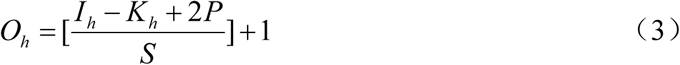

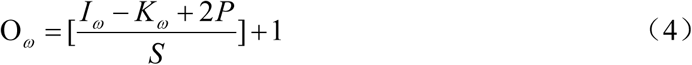

Where *O_h_* and *O_ω_* denote the output height and output width, respectively; *I_h_* and *I_ω_* represent the input height and input width, respectively;*K_h_* and *K_ω_* are the kernel height and kernel width, respectively; *P* is the padding size; and *S* is the stride size. Average pooling is adopted to downsample the time dimension, which preserves information from key time points while reducing computational complexity and adapting to the global feature learning of 10-second-long data blocks.

(3) Training Optimization Module: The function of this module is to achieve efficient model convergence and optimize the model’s performance on EEG signal data through scientific training strategies, with its flowchart illustrated in Figure 2. In this module, Focal Loss is used to dynamically adjust class weights and focus on hard-to-classify epileptic samples. For binary classification (normal vs. epileptic signals), the Focal Loss function is defined as follows**^Error! Reference source not found.^**:

**Figure 2.**
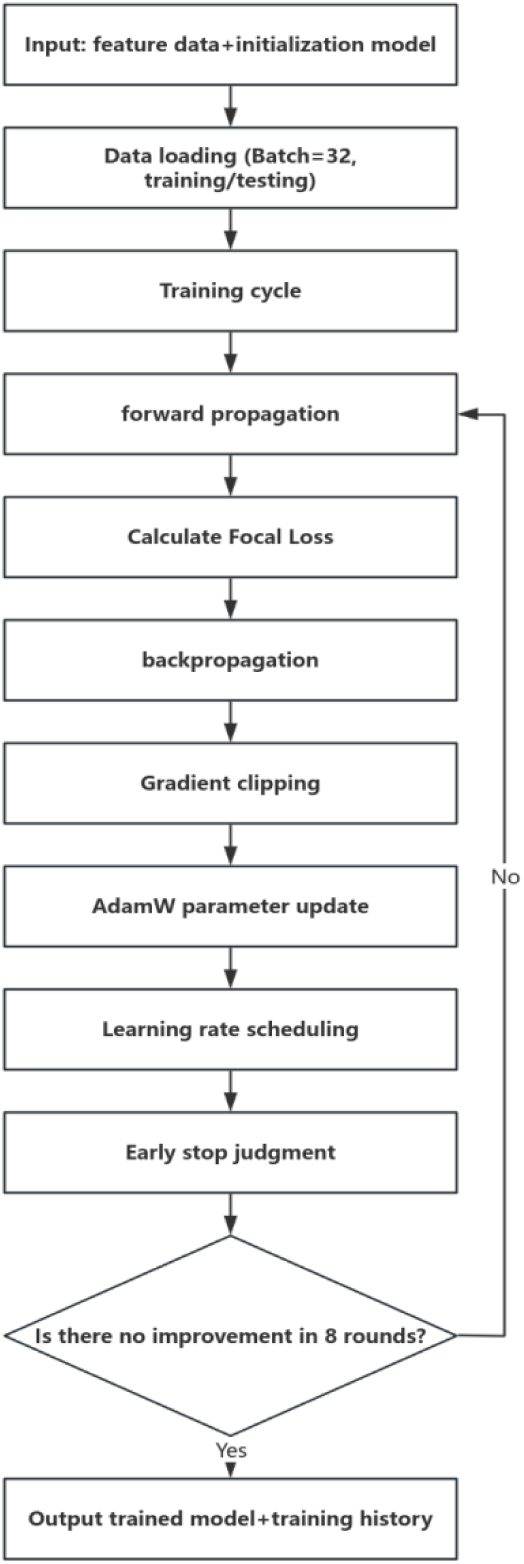
Training Optimization Flow Chart

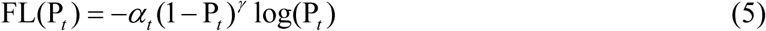

where *P_t_* is the predicted probability for the true class; *α_t_* balances class imbalance; and *γ* emphasizes hard samples. The Adam optimizer AdamW optimizer is selected, combined with L2 regularization, to accelerate convergence and suppress overfitting.

For the training process, an early stopping mechanism is adopted—if there is no performance improvement for 8 consecutive epochs, training will automatically terminate after these 8 epochs. When the loss stagnates, the learning rate is reduced to 0.3 times its original value. To prevent gradient explosion, gradient clipping is applied, and 5-fold cross-validation is used to ensure the stability and reliability of the model performance.

(4) Epilepsy Detection and Evaluation Module: The function of this module is to evaluate the model’s ability to detect epileptic EEG signals and verify the system’s practicality. The optimal classification threshold within the range of 0.3–0.7 is searched for to balance the clinical requirements for misdiagnosis and missed diagnosis rates. Core metrics for evaluating classification performance—accuracy, recall, and precision—are then calculated. For binary classification tasks, these metrics are defined as follows^[25]^:

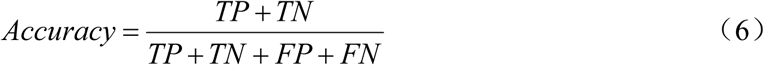

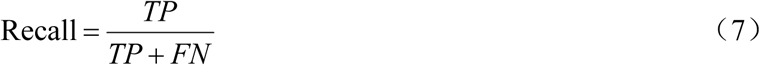

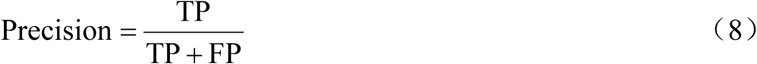

where *TP* (True Positive) denotes the number of correctly detected epileptic signals, *TN* (True Negative) represents the number of correctly detected normal signals, *FP* (False Positive) refers to the number of normal signals misdiagnosed as epileptic, and *FN* (False Negative) indicates the number of epileptic signals missed (i.e., misdiagnosed as normal). Confusion matrices and classification reports are generated to visually illustrate the model’s recognition performance on normal and epileptic EEG signals. Finally, the mean and standard deviation of the 5-fold cross-validation results are calculated to verify the system’s stability.

## 4. Experiment and Analysis

### 4.1 Experimental results

In this section, the experiment evaluates the performance of three different models—EEGNet, AlexNet, and ShallowConvNet—after training, where features for the models are extracted using Continuous Wavelet Transform (CWT) and Short-Time Fourier Transform (STFT) respectively. For the performance evaluation of the deep learning models in this experiment, three key metrics are selected: accuracy, precision, and recall. The performance metrics obtained from the experiment are presented in Table 1 and Table 2, and the confusion matrices are illustrated in Figure 3 and Figure 4. The ratio of normal samples to abnormal samples in the confusion matrices is approximately 2:1.

**Figure 3:**
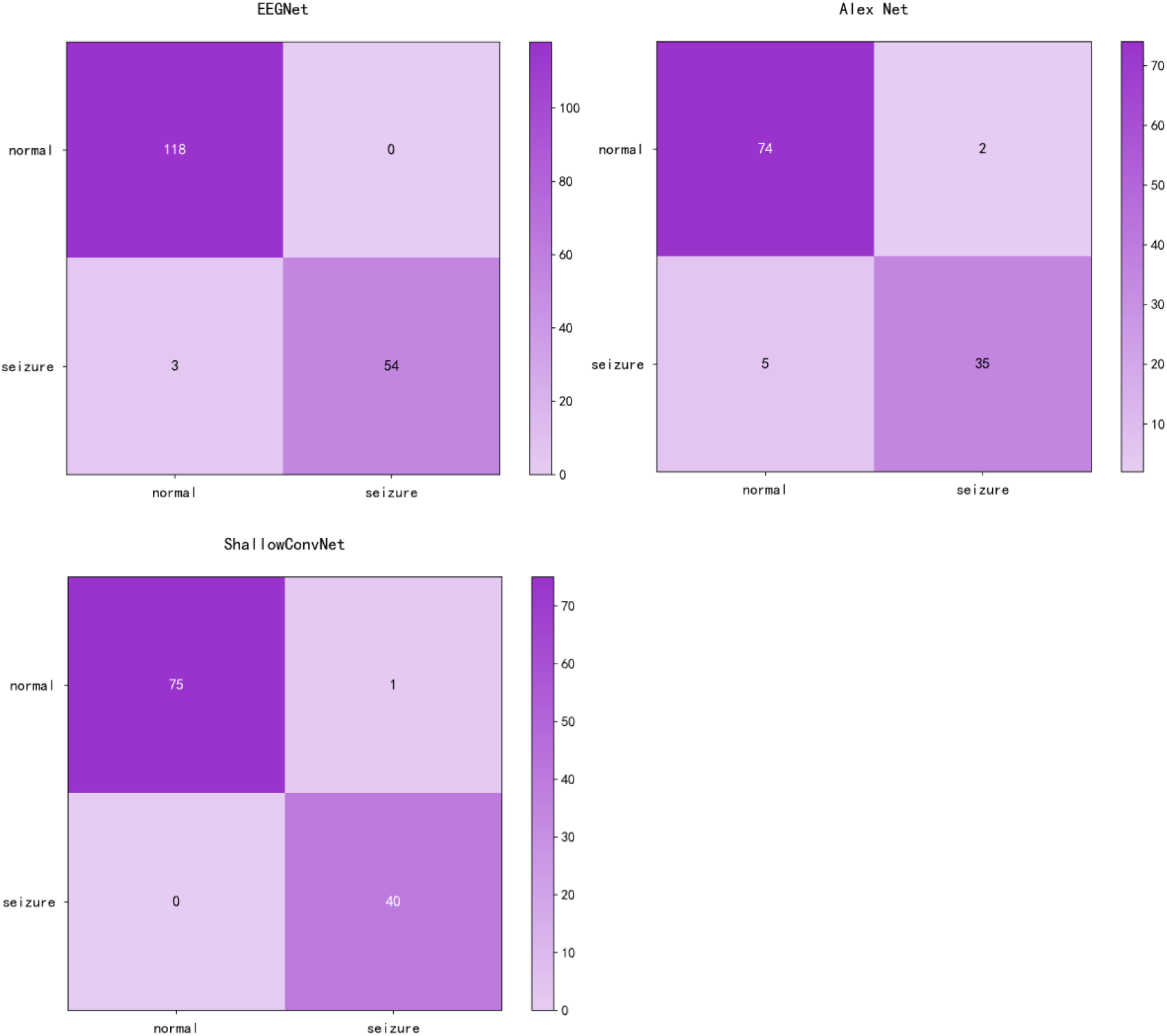
Confusion Matrix Using CWT

**Figure 4:**
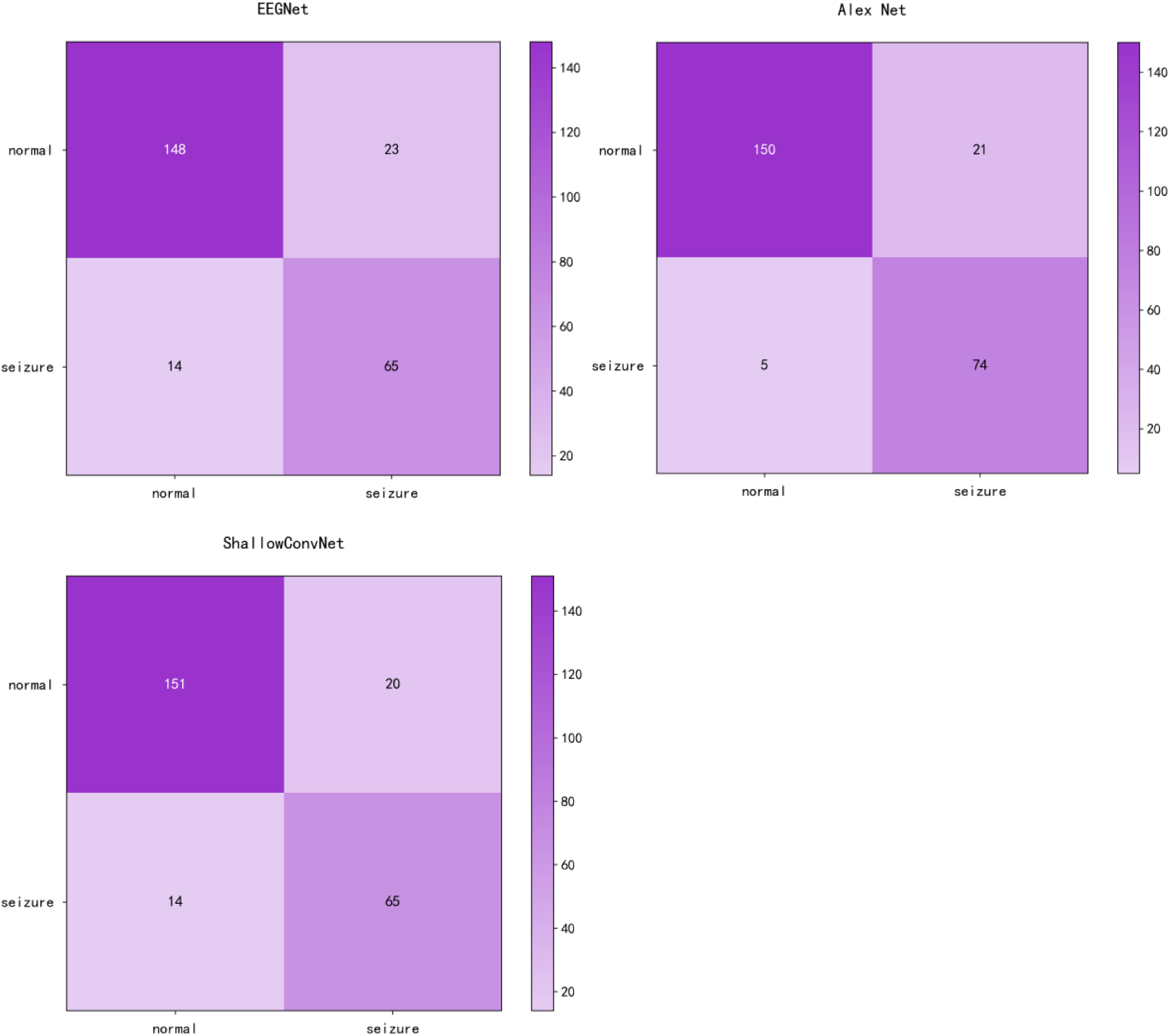
Confusion Matrix Using STFT

**Table 1.** Performance indicators using CWT.

| Model | Accuracy (%) | Precision(%) | Recall(%) |
| --- | --- | --- | --- |
| CWT+EEGNet | 98.29 | 100 | 94.74 |
| CWT+AlexNet | 93.97 | 94.59 | 87.50 |
| CWT+ShallowConvNet | 99.14 | 97.56 | 100 |

**Table 2.** Performance indicators using STFT.

| Model | Accuracy (%) | Precision(%) | Recall(%) |
| --- | --- | --- | --- |
| STFT+EEGNet | 85.20 | 73.86 | 82.28 |
| STFT+AlexNet | 89.60 | 77.89 | 93.67 |
| STFT+ShallowConvNet | 86.40 | 76.47 | 82.28 |

According to Tables 1 and 2, the combination of feature extraction methods and models has a significant impact on classification performance. The accuracy of the experimental groups using CWT exceeded 90%, which was overall better than that of the STFT experimental groups (with an accuracy range of 80%–90%). Among these combinations, the CWT+ShallowConvNet pair achieved the highest accuracy of 99.14%, closely followed by CWT+EEGNet at 98.29%. In terms of precision, CWT+ShallowConvNet reached 97.56%, while CWT+EEGNet achieved 100%—the latter was superior in reducing false alarms for abnormal signals. Regarding recall, when using the EEGNet and ShallowConvNet models, feature extraction with CWT yielded better performance than the STFT experimental groups; in contrast, the opposite was true when using the AlexNet model. Notably, CWT+ShallowConvNet achieved a recall of 100%, indicating the strongest ability to detect abnormal signals. As observed from the confusion matrices in Figures 3 and 4, the system adopting the combination of CWT and the ShallowConvNet model exhibited the best performance.

### 4.2 Comparison of time-frequency characteristics

To visually compare the performance of Short-Time Fourier Transform (STFT) and Continuous Wavelet Transform (CWT) in extracting features from normal and epileptic EEG signals, EEG signals from the FP1-F7 channel during both normal periods and epileptic seizure periods were selected (as shown in Figure 5). These signals were then subjected to the two transforms, and the results were visualized. The visualization outcomes are presented in Figures 6 and 7.

**Figure 5:**
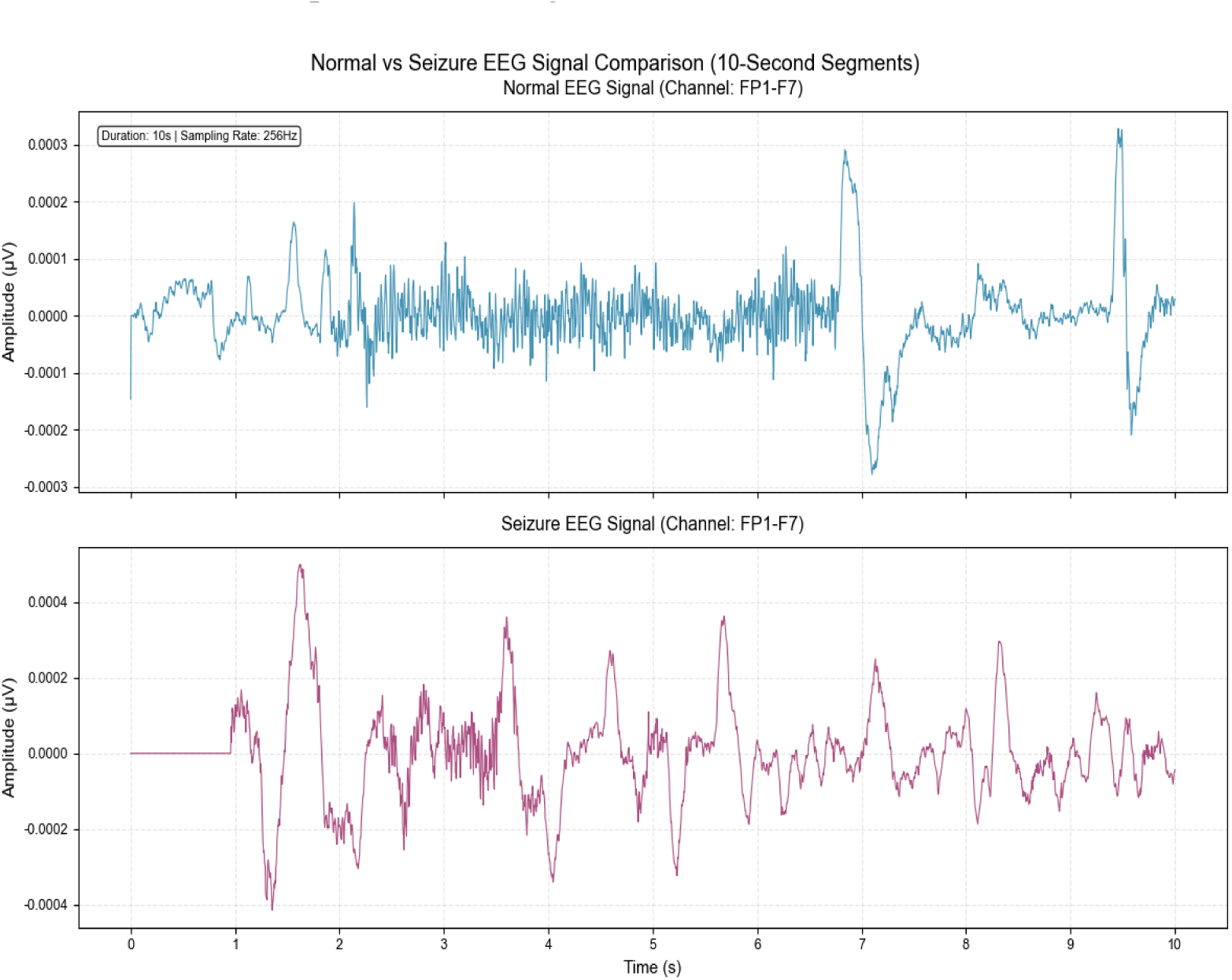
Original Signal Diagram of Normal EEG and Epilepsy EEG

**Figure 6.**
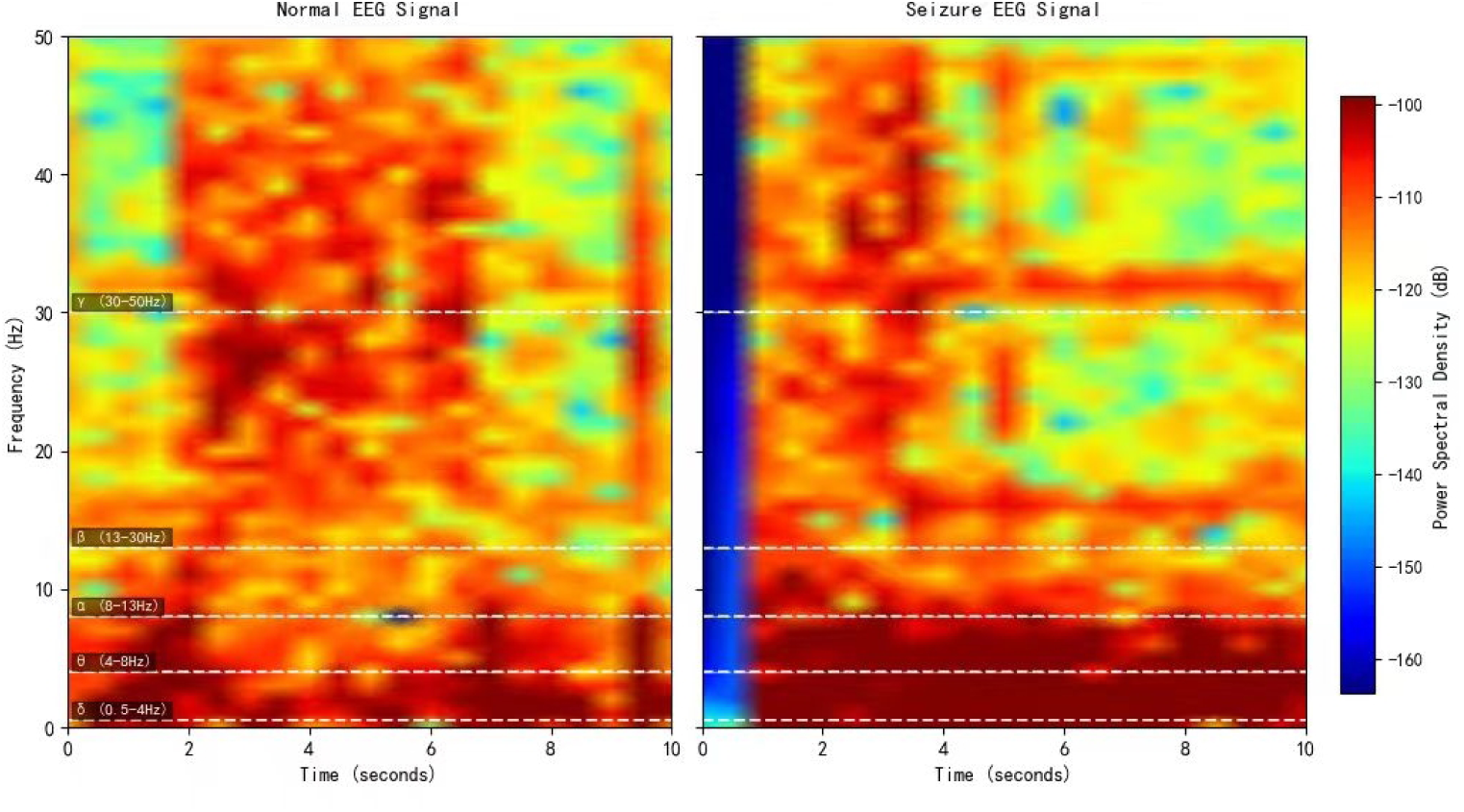
STFT time-frequency comparison chart

**Figure 7.**
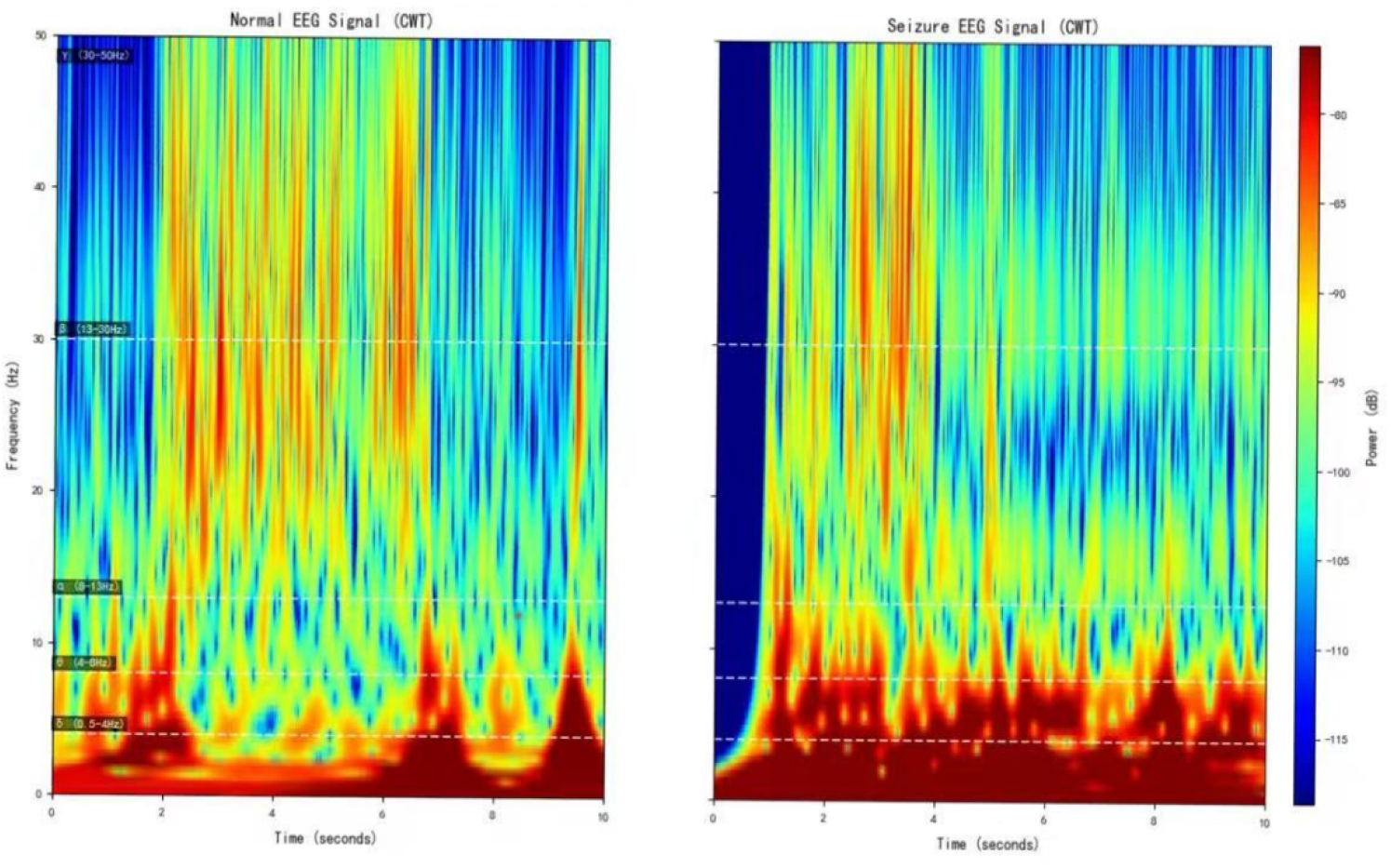
CWT time-frequency comparison chart

As observed in Figure 6, the energy distribution of normal EEG signals across different frequency bands exhibits fuzzy and fragmented characteristics. Additionally, due to the “averaging effect” of the fixed time window in STFT, the abnormal energy changes corresponding to epileptic EEG signals during seizure periods cannot be accurately localized. In contrast, by analyzing Figure 7, it can be seen that the energy distribution of normal EEG signals shows clear hierarchical continuity. For epileptic EEG signals (corresponding to seizures), distinct pathological features—such as spike waves and sharp waves—are observed, and the precise localization of energy changes in each frequency band along the time dimension enables clear tracing of the time-frequency evolution process of abnormal discharges.

From these observations, it is evident that the time-frequency analysis mechanism of CWT is more aligned with the non-stationary and multi-feature coupling characteristics of epileptic EEG signals. The time-frequency maps generated by CWT retain richer and more accurate pathological features, laying a foundation for the efficient classification of subsequent deep learning models. Therefore, CWT demonstrates stronger adaptability in the EEG signal processing of this experiment.

## 5 Conclusions

This experiment focuses on the classification task of epileptic EEG signals and conducts an in-depth comparison of the adaptability of different time-frequency transformation methods and deep learning models. The research findings indicate that the experimental group using Continuous Wavelet Transform (CWT) exhibits significantly better performance in preserving the pathological features of normal and epileptic EEG signals compared to the group using Short-Time Fourier Transform (STFT). Specifically, CWT can effectively process temporal information and clearly distinguish the energy distribution across the alpha, theta, beta, delta, and gamma frequency bands. In contrast, STFT suffers from a “smoothing effect” caused by its fixed time window, which results in blurred and diffused signal energy, thereby weakening feature recognition. Consequently, the time-frequency maps generated by CWT provide higher-quality input for subsequent models, and their core performance metrics are comprehensively superior to those of the STFT-based scheme.

When using CWT-extracted features as input, the performance of different models varies:

(1) ShallowConvNet directly captures features using shallow large convolutional kernels, achieving a recall of 100% and an accuracy of 99.14%. Owing to its lightweight structure, the model also maintains a precision of 97.56%.
(2) EEGNet leverages depthwise separable convolution and an SE (Squeeze-and-Excitation) attention mechanism to suppress noise, attaining a precision of 100% and demonstrating a stronger ability to reduce false alarms for abnormal signals.

Based on the research results, future work will be advanced in four directions:

(1) Construct heterogeneous datasets while exploring multi-channel spatial features. This involves integrating multi-center data to verify the model’s generalization ability and designing cross-channel modules to excavate the spatial propagation patterns of abnormal discharges.
(2) Explore learnable time-frequency transformation techniques, particularly end-to-end learnable wavelet transform, to achieve collaborative optimization of time-frequency parameters and model parameters.
(3) Enhance model interpretability by utilizing activation maps and attention visualization to analyze the model’ s focus regions, thereby improving its clinical credibility.
(4) Promote lightweight deployment by designing a mobile solution based on ShallowConvNet, verifying the real-time performance of wearable devices, and facilitating the transition of this technology from laboratory research to clinical auxiliary diagnosis.

## Data Availability

All EEG data used in this study are from the CHB-MIT Scalp EEG Database, which is publicly available at https://physionet.org/content/chbmit/1.0.0/ (accession details: dataset name “CHB-MIT”, available without restriction).

## Acknowledgments

The authors sincerely thank the Children’s Hospital Boston (CHB) and Massachusetts Institute of Technology (MIT) for making the CHB-MIT Scalp EEG Database (CHB dataset) publicly available, as this high-quality annotated EEG data laid a solid foundation for our epilepsy automatic detection research.

This work was supported by the Young Academic and Technical Leaders Program of Yunnan Province (Grant No.: 202305AC160077) and the Scientific Research Fund of the Yunnan Provincial Department of Education (Grant No.: 2025Y0667). We also acknowledge support from the School of Electrical and Information Engineering, Yunnan Minzu University、Yunnan Key Laboratory of Unmanned Autonomous System and Key Laboratory of Cyber-Physical Power System of Yunnan Colleges and Universities for providing research resources.

## Author Contributions Conceptualization

Paul Nuyujukian. Data curation:Canhui Wang.

Formal analysis: Canhui Wang,Haoran Tang,Tianqi Xu.

Funding acquisition: Yan Li.

Investigation:Canhui Wang, Zongfang Ren.

Methodology:Canhui Wang.

Project administration:Canhui Wang,Yan Li.

Resources: Yan Li

